# Analysis of epidemiological characteristics of coronavirus 2019 infection and preventive measures in Shenzhen China—a heavy population city

**DOI:** 10.1101/2020.02.28.20028555

**Authors:** Kai Yang, Lingwei Wang, Furong Li, Dandan Chen, Xi Li, Chen Qiu, Rongchang Chen

**Author notes:** Correspondence: Rongchang Chen,; Chen Qiu. Kai Yang and Lingwei Wang contributed equally to this work.

## Abstract

Coronavirus 2019 infection (COVID-19) outbroke in Wuhan, Hubei and spread to all provinces in China and other countries. Shenzhen ranked the top cities outside Wuhan with reported 416 confirmed cases by February 20, 2020. Here, we analyzed the epidemiological characteristics of COVID-19 in Shenzhen and potential link to the preventive strategies for the whole city and inside hospitals. Based on the daily new cases, the epidemic of COVID-19 in Shenzhen can be classified into three phases: the slow increase phase from January 19 to January 28, the rapid increase and plateau phase from January 29 to February 5 and the decline phase since February 6. In the three phases, the number of patients from Hubei decreased, and the number of familial clustering cases increased. The newly diagnosed COVID-19 cases reached its peak around January 31, which was 7 days after the peak date of cases arrival at Shenzhen. A series of early preventive strategies were implemented since January 19, which included detection of body temperature at all entrances of main traffic and buildings, outpatients service specially for patients with fever in all main hospitals in Shenzhen. All the patients with fever were screened with nasal or throat swab PCR detection of coronavirus 2019, Chest CT and blood lymphocyte counting in order to find out early case of COVID-19. Observation wards were established in every main hospital and a designated hospital was responsible for admission and medical care of all confirmed cases. Protection procedure was established for all medical staff involved in the screening and care of suspected and confirmed cases. 14 days isolated observation of all subjects arrived at Shenzhen from Hubei was implemented in February 2. After the implementation of all these strategies and measures, the COVID-19 cases started to decline since February 6. There were almost no community transmission and nosocomial infection occurred in Shenzhen.

In conclusion, in situation of major outbreak of respiratory infectious disease, such as COVID-19, in nearby province of Hubei, Shenzhen, a high population density, high proportion of external population and high mobility city, has to face the imported cases and risk of spreading the outbreak into Shenzhen city. The implementation of early preventive strategies and measures in Shenzhen were successful in early identification of COVID-19 cases and prevented major outbreak occurred in Shenzhen. Early identification of imported cases, prevention of family clustering transmission, preventive measures in the public area and very strict infection control procedure in hospital setting are crucial for the successful control of outbreak in Shenzhen.

## Introduction

Novel coronavirus pneumonia (COVID-19) patients were firstly reported in Wuhan, Hubei in December 2019, and spread to all provinces in China and more than 20 countries in the following month[1-5]. By February 20, 2020, COVID-19 has infected more than 70000 cases and led to more than 2000 deaths[6]. Based on the announcement of National Health Commission of People’s Republic of China by February 20, 2020, Shenzhen ranked the top cities outside Wuhan with reported 416 confirmed cases. There were some cities with heavy population density and migrant workers, such as Shenzhen, Beijing, Shanghai and Guangzhou. There are about 20 million people living in Shenzhen, of which the external population from the internal area of China, including Hubei province, account for a large proportion. High population density, high proportion of external population and high mobility may increase the possibility of COVID-19 outbreak[7, 8]. The first COVID-19 patient in Shenzhen was admitted on January 9, 2020, and 416 cases have been confirmed by February 20, 2020. Most of the confirmed cases were imported cases from Hubei province until now. There was no large-scale transmission and nosocomial infection in Shenzhen city so far. Therefore, the purpose of the study was to analyze the epidemiology and preventive strategies in Shenzhen in order to understand the main transmission route and effective preventive strategies in cities with risk of imported cases, which may provide clue for better preventing outbreak of potential respiratory infectious disease, such as COVID-19 in cities with heavy population density and high proportion of external population.

## Method

The data of this study were downloaded from the data open-platform of Shenzhen government (https://opendata.sz.gov.cn). The dynamic change of accumulated confirmed cases and newly diagnosis of each day were used for the analysis. The diagnosis criteria of confirmed cases in Shenzhen was consistent with the consensus of diagnosis and management of COVID-19 by National Health Commission of People’s republic of China[9]. History of visit of Hubei was defined as those who live in Hubei and came to Shenzhen, or who live in Shenzhen but recently visited Hubei and returned within 14 days before the onset of symptoms related to COVID-19. History of contact with Hubei patients was defined as those who contacted with COVID-19 patients from Hubei within 14 days before the onset. History of contact with non-Hubei COVID-19 patients was defined as those who contacted with COVID-19 patients from other areas except Hubei within 14 days before the onset. No contact history was defined as those who denied contact with any other COVID-19 patients. Familial clustering was defined as 2 or more patients from the same family. Hospital environment transmission was defined as COVID-19 related to exposure inside hospital, including staff working in hospital, subjects who admitted into hospital due to non-COVID-19 disorders, or visited to hospital due to other reasons within 14 days before the COVID-19 onset. Other definitions were consistent with previous studies[10, 11]. Analysis of variance or Chi square test were used for the comparison of 3 groups. Statistical analysis was performed in SAS 9.4 (SAS Inc, Cary, NC, USA), and P<0.05 was considered as statistically significant.

## Results

### 1. Trend of COVID-19 incidence in Shenzhen

The first COVID-19 patient in Shenzhen was admitted on January 9, 2020. The number of confirmed cases of COVID-19 began to be released daily on January 19, 2020. Based on the daily new cases, three phases were classified. The slow increase phase begun from January 19 to January 28, the rapid increase and plateau phase begun from January 29 to February 5 and the decline phase begun since February 6 (Figure 1). For those confirmed cases, the exact date of arriving at Shenzhen were identified. The peak arrival time was around January 23, which was 7 days on average ahead of the peak of daily new confirmed cases (Figures 1A and 1C).

**Figure 1.**
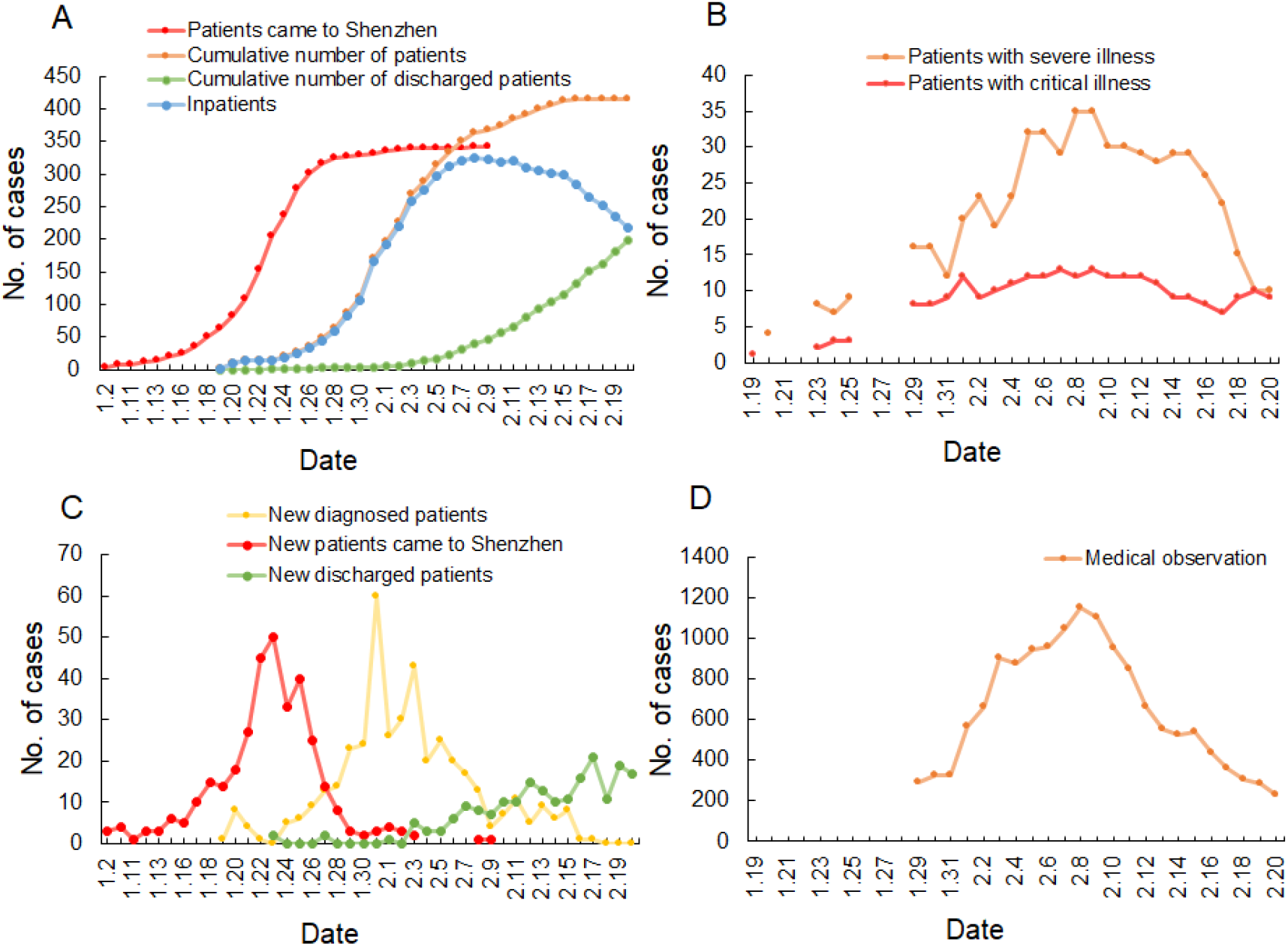
Epidemic curve of COVID-19 in Shenzhen by February 20, 2020. (A) Cumulative number of patients for different indexes. (B)Patients with severe and critical illness. (C) The trend of newly patients came to Shenzhen and newly diagnosed patients. (D) Patients with medical observation.

### 2. Prevention of COVID-19 in Shenzhen

A series of preventive strategies and measures were shown in Fig 2. We divided the preventive strategies into early prevention and strengthened prevention.

**Figure 2.**
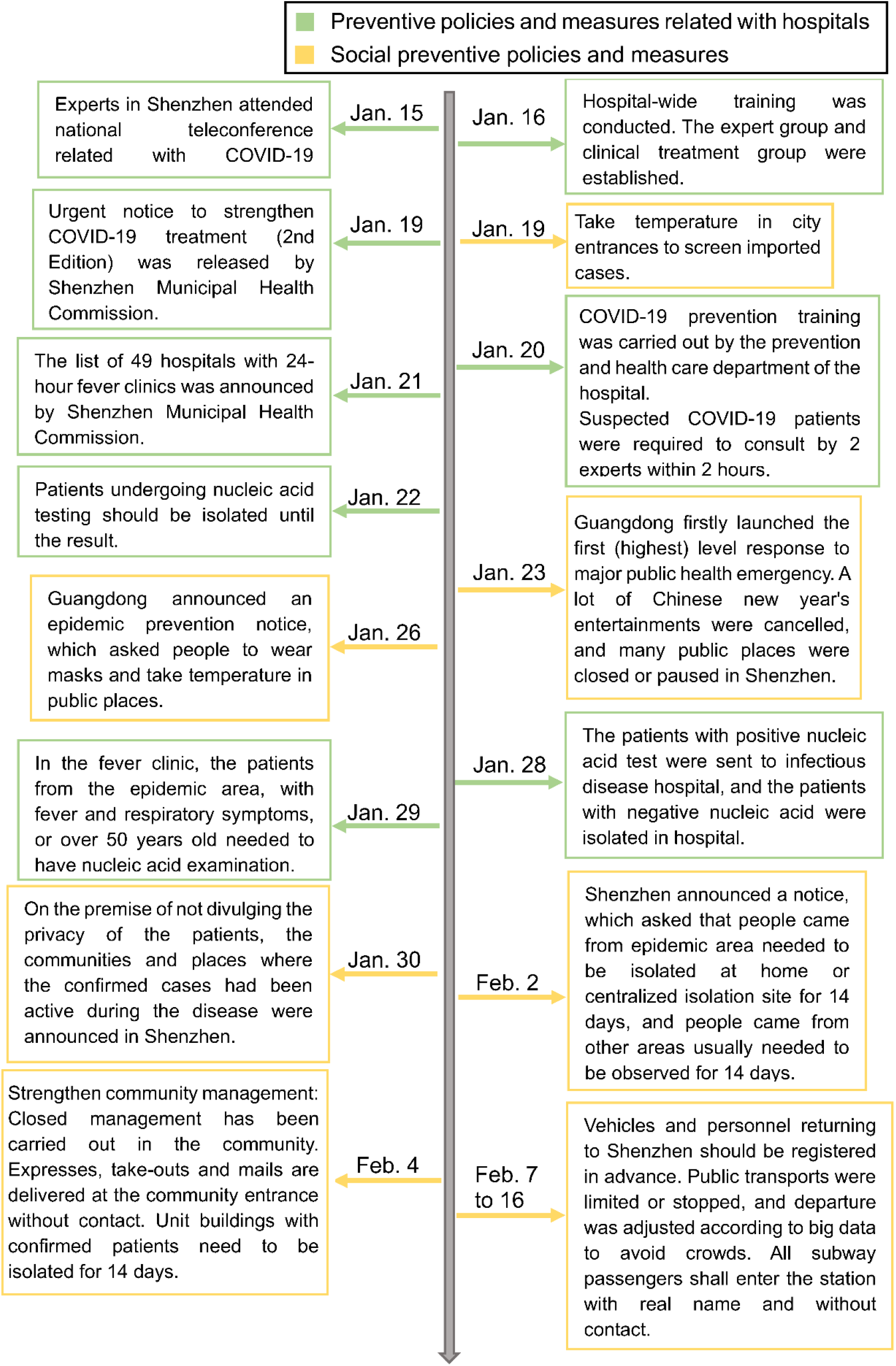
Main preventive policies and strategies of COVID-19 in Shenzhen, which could be found in the website of National Health Commission of China (http://www.nhc.gov.cn/), Health Commission of Guangdong Province (http://wsjkw.gd.gov.cn/), and Shenzhen Municipal Health Commission (http://wjw.sz.gov.cn/).

#### 2.1 Early preventive policies and strategies

Early preventive strategies and measures against COVID-19 included public announcement of the outbreak in Wuhan, Hubei and preparation for the diagnosis and management of COVID-19 in hospitals (Figure 2).

The hospitals in Shenzhen have begun to prepare for the possible outbreak of COVID-19 since January 15, including the training of medical staff and epidemiological investigators, hospital disinfection, medical material reserve, outpatient adjustment, medical waste management, and so on. All 49 hospitals in Shenzhen set up 24-hour fever clinics for screening of possible COVID-19 with nasal or throat swab PCR detection of coronavirus 2019, Chest CT and blood lymphocyte counting. Suspected patients would be isolated and observed in single room for each case with special preparation of infection control measures in the hospital. Confirmed patients will be transferred to the Second Affiliated Hospital of South University of Science and Technology (Shenzhen infectious disease hospital) for treatment with negative-pressure ambulance, which was a designated hospital for inpatient management of infectious disease.

All medical staff involved in the management of COVID-19 patients implemented infection control procedures, which included work clothes, disposable work caps, disposable gloves, disposable medical protective clothing, medical protective masks or powered air supply filter respirators, protective screens or goggles, work shoes or rubber boots, waterproof boot covers.

For the general population, people entering the hospital were asked to wear masks and take body temperature. The hospital environment was disinfected timely and regularly, hand-washing equipment and disinfectants were equipped in multiple places, and indoor air ventilation was strengthened.

As early as January 19, when the National Health Commission announced the first imported case in Shenzhen, Shenzhen began to take temperature for people in main city entrances to screen imported cases. After the release of the first (highest) level response to major public health emergencies in Guangdong on January 23, all Chinese new year’s entertainments were cancelled, and many public places were temporarily closed in Shenzhen, including market, cinema, museum, library, gymnasium, and so on. Other necessary public places needed to be disinfected regularly, including airport, station, port, freeway entrance, urban traffic, community entrance. The employees in these places needed to have health examinations. People came to these places needed to wear masks and take body temperature.

#### 2.2 Strengthened the preventive measures in Shenzhen

Millions of people returned to Shenzhen from all over the country after Chinese new year’s holiday, which may lead to the imported cases and potential transmission. Therefore, strengthened measures were implemented in February 2, which included isolating all the new arrival people came from epidemic area for medical observation for 14 days, informing all people in the living communities once there were confirmed cases ever stayed in the communities, using of big data and information technology to track the cities of the subject had visited in the last 14 days and identify the contacted persons of the confirmed cases (Figure 2).

#### 2.3 The relationship analysis between the new confirmed cases and preventive strategies

As transmission of infectious disease requires links between three parts of the cycle, the source of infection, route of transmission and susceptible population. The preventive strategies and measures implemented in Shenzhen were supposed to block the links by early identifying and isolating potential and confirmed cases as well as cutting of the transmission route. The newly diagnosed COVID-19 cases reached its peak around January 31, which was 7 days after the peak date of cases arrival at Shenzhen and around 10 days after the serial early preventive strategy implemented. Taking account of the incubation period (mostly 3-7 days, with mean of 3.7 days) and the time between symptom onset and confirm of the diagnosis (6 day on average) [9, 12], the peak of new confirmed cases coincided with the implementation of serial preventive strategy and measures,indicating these preventive strategies and measures were effective in preventing transmission of COVID-19 in Shenzhen.

### 3. Epidemiological characteristics of COVID-19 in Shenzhen

#### 3.1 Patient characteristics and route of transmission

Patients demographic data were shown in Tab. 1. Patients were divided into 3 groups according to the time phase of COVID-19 incidence. Compared with the reports from Wuhan, Hubei[11], the age of infected population in Shenzhen was younger and decreasing gradually, in which 33 patients were children. There was no difference in gender (Table 1).

**Table 1.** Demographic and clinical characteristics in of COVID-19 patients.

| **Parameters** | **1.19-1.28** | **1.29-2.5** | **2.5-2.20** | **$F/\chi^2$** | ***P*** |
| --- | --- | --- | --- | --- | --- |
| **N** | 63 | 228 | 125 |  |  |
| **Age** |  |  |  |  |  |
| Mean±SD | 51.8±17.0 | 45.3±17.5 | 42.4±17.5 | 6.10 | 0.002 |
| Median (Min, Max) | 56(7,78) | 47(1,86) | 42(2,72) |  |  |
| **Gender (Male)** | 34(54.0) | 104(45.6) | 60(48.0) | 1.39 | 0.498 |
| **Infection pattern** |  |  |  |  |  |
| From Hubei | 61(96.8) | 174(76.7) | 74(59.2) | 34.47 | <0.001 |
| Contact with Hubei patients | 2(3.2) | 27(11.9) | 27(21.6) |  |  |
| Contact with non-Hubei patients | 0(0.0) | 12(5.3) | 15(12.0) |  |  |
| No contact | 0(0.0) | 14(6.2) | 9(7.2) |  |  |
| **Familial clustering** |  |  |  |  |  |
| Yes | 24(38.1) | 132(57.9) | 76(61.3) | 9.93 | 0.007 |
| No | 39(61.9) | 96(42.1) | 48(38.7) |  |  |

There were 63 cases in slow increase phase from January 19 to January 28. Almost all cases (61/63, 96.8%) were patients from Hubei, and 2 cases were family members of patients from Hubei, indicating that main source of COVID-19 patients was imported from Hubei province at this beginning slow increase phase.

In the rapid increase and plateau phase from January 19 to February 5, there were 228 confirmed patients. Compared with the previous period, although majority of the patients (174, 76.7%) were still from Hubei, but infection related to contact with Hubei patients (27, 11.9%), non-Hubei patients (12, 5.3%) and no contact history (14, 6.2%) increased. Meanwhile, cases of familial clustering increased by 19.8%, indicating that local transmission in the family environment should not be ignored.

In the decline phase since February 6, although the number of daily new confirmed cases decreased significantly, but infection related to contact with Hubei patients (27, 21.6%), non-Hubei patients (15, 12.0%) and no contact history (9, 7.2%) had the tendency of increase. Meanwhile, cases of familial clustering increased to be predominant (76, 61.3%), indicating that local transmission in the family environment was important route of transmission.

#### 3.2 Familial clustering of COVID-19 in Shenzhen

Familial clustering was an important feature of COVID-19[10]. In Shenzhen, 232 COVID-19 patients (232/416, 55.8%) were come from 86 families, 15 of which had more than 3 patients (Figure 4). Patients in the same family may be infected by other patients simultaneously or by other family members.

**Figure 3.**
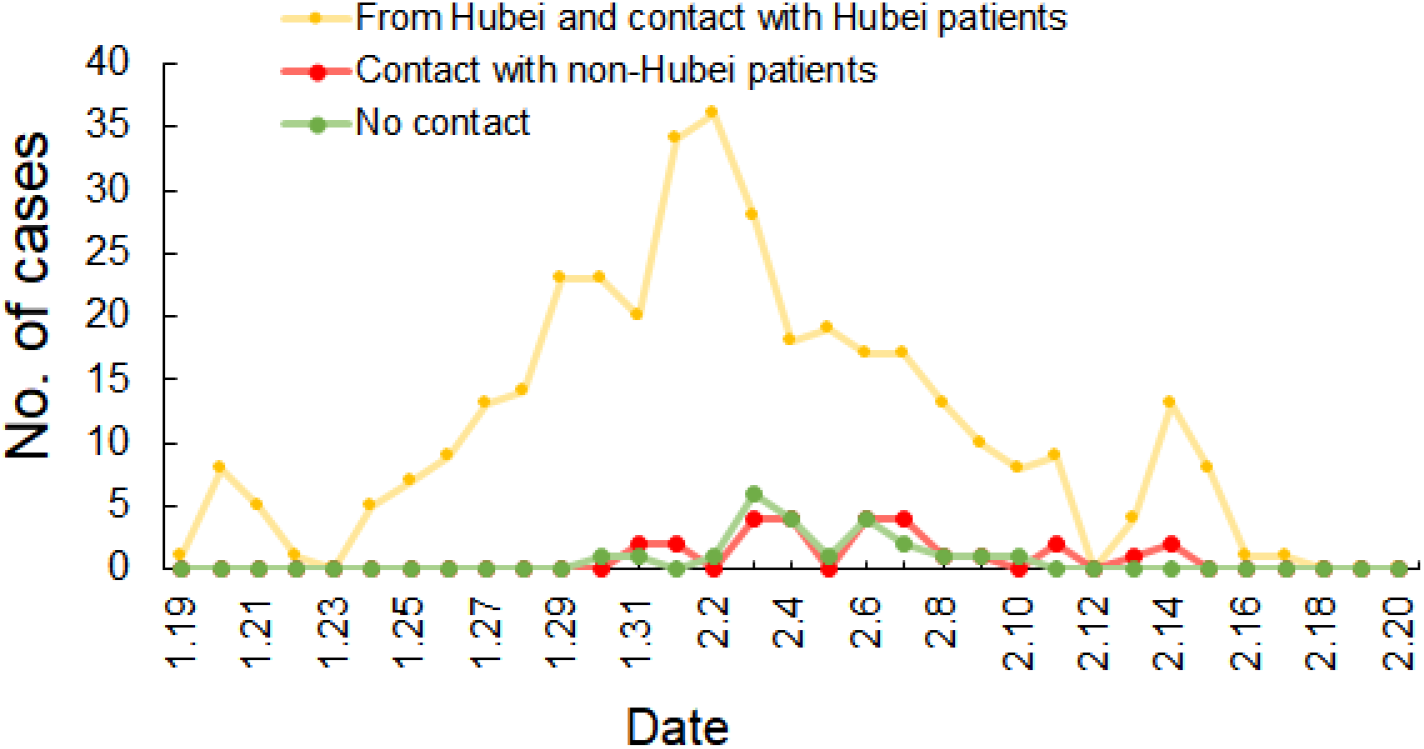
Epidemic curve of COVID-19 patients with different infectious pattern in Shenzhen by February 20, 2020.

**Figure 4.**
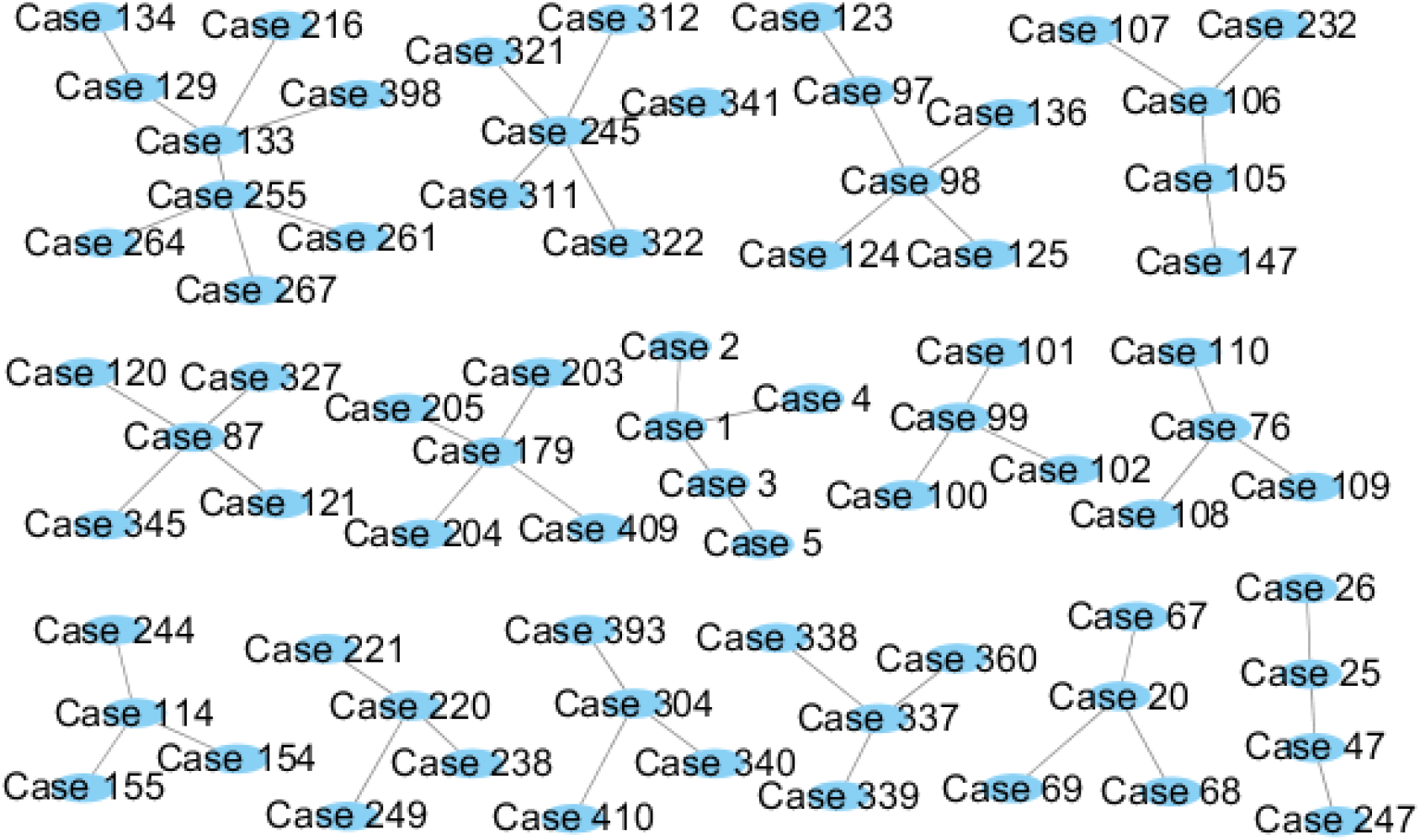
Infection chain of family clustering cases with more than 4 people.

### 4. The transmission in hospital environment

There were reports from Wuhan, Hubei where the pandemic started, infection acquired in hospital environment accounted up to 41.3% of the COVID-19 patients [12]. Subjects infected in hospital environment included medical staff and patients admitted into hospitalized for other reasons. In comparison with those reports from Wuhan, Hubei, there was no infection among medical staff or patients infected in hospital environment in Shenzhen hospitals until February 20.

## Discussion

The pandemic of COVID-19 started in Wuhan, Hubei, China in December 2019 and has spread throughout China and many countries worldwide. Based on the data release from the National Health Commission of People’s Republic of China by February 20, 2020, Shenzhen ranked the top cities outside Wuhan with reported 416 confirmed cases. However, the outbreak in Shenzhen was under control so far in a relatively short time with limited cases with regard to its population density. Among the 416 confirmed cases, 87.7% of the COVID-19 patients were imported cases from Hubei or patients contacted with families from Hubei. Only a few of patients were probably caused by community transmission or contact with other people in Shenzhen. Through the data analysis, the implementation of early preventive strategies and measures coincided with peak of daily new confirmed cases and started to decline afterward. The results strongly supported that the early preventive strategies and measures were effective in preventing the outbreak of COVID-19 in Shenzhen.

Shenzhen, Guangdong province located in the south of China, has several important factors susceptible to the spread of COVID-19 into the city. There are about 20 million people living in Shenzhen, of which the external population from the internal area of China, including Hubei province, account for a large proportion. High population density, high proportion of external population and high mobility may increase the possibility of COVID-19 outbreak[7, 8]. In addition, the time around Chinese New Year (January 25) was the peak time period for people to travel from the coastal cities to inland cities or backward. All these factors put Shenzhen into the high risk of outbreak.

Through the analysis, it was revealed that majority of the confirmed cases was imported cases from Hubei. Families cluster was another important characteristic of the patients. Subjects in incubation or early stage of illness (with minimal symptoms) who came to Shenzhen and lived together or closely contacted with relatives or friends (such as having dinner together), were important route of infection in Shenzhen. These cases demonstrated the need to reduce gatherings.

The early preventive strategies and measures in Shenzhen focused on the early identification and isolation of COVID-19 patients (the main source of infection) and cutting off the route of transmission. For cities with risk of imported cases, population inflow was the most important factor of transmission of infectious diseases. Therefore, screening suspected patients and persons from epidemic areas in city entrances, and isolation according to the incubation period can find possible sources of infection early and prevent the spread of infection diseases, which were also widely used in the prevention of other respiratory infectious diseases, such as influenza in United States and SARS in China[13, 14]. Protective measures for the general population, such as disinfection, wearing masks, measuring body temperature, preventing crowd gathering and reducing human contact, can also prevented the spread of respiratory infectious diseases[15, 16]. For staff in public places, protection measures were both important for the public people and themselves. Information publicity also played an important role in the prevention of respiratory infectious diseases for residents[17]. Big data and artificial intelligence were increasingly important in preventing infectious, which can monitor the population and formulate corresponding measures in time to reduce the spread of the respiratory infectious disease, and easily track patients and close contacts[18].

Hospitals and medical staff played the most important role in the treatment of respiratory infectious diseases. Due to the progression of the illness and many patients seeking medical care in the hospital, human-to-human infection was more likely to occur than other places, so more strict protections should be taken[19, 20]. The medical staff, especially those in infectious disease department, emergency department, respiratory department and other departments related with infectious disease prevention, should have the ability to discern patients with respiratory infectious diseases and take protective measures[21-23]. When there are warning of respiratory infectious diseases, hospitals needed to take measures to prepare for medical staff training, protective measures and medical supplies earlier than other places[24]. Meanwhile, it is necessary to isolate patients with infectious diseases and suspected patients away from other patients to avoid nosocomial infection[25]. In conclusion, in situation of major outbreak of respiratory infectious disease, such as COVID-19, in nearby province of Hubei, Shenzhen, a high population density, high proportion of external population and high mobility city, has to face the imported cases and risk of spreading the outbreak into Shenzhen city. The implementation of early preventive strategies and measures in Shenzhen were successful in early identification of COVID-19 cases and prevented major outbreak occurred in Shenzhen. Early identification of imported cases, prevention of family clustering transmission, preventive measures in the public area and very strict infection control procedure in hospital setting are crucial for the successful control of outbreak in Shenzhen.

## Data Availability

Data were released in the website of data open platform of Shenzhen Municipal Government.

https://opendata.sz.gov.cn

## Conflict of Interest

No potential conflicts of interest were disclosed.

## Funding

This work was supported by emergency research project of coronavirus 2019 infection (No. 2020XGZX024).

## Notes

### Competing Interest Statement

The authors have declared no competing interest.

